# A systems approach to enhance Lynch syndrome diagnosis through tumor testing

**DOI:** 10.1101/2022.06.13.22276231

**Authors:** Vinit Singh, Catherine Mezzacappa, Peter Gershkovich, Jessica Di Giovanna, Amanda Ganzak, Joanna Gibson, John Sinard, Rosa M. Xicola, Xavier Llor

## Abstract

**Background:** Guidelines recommend universal mismatch repair tumor (MMR) testing of colorectal adenocarcinomas (CRC) to screen for Lynch syndrome (LS). However, its implementation remains disjointed and referral for genetic testing dismal, particularly among minorities. We aimed to increase referral, cancer genetic testing, and eventually LS diagnosis by developing a systems approach which, in the second phase was automated.

**Methods:** This is a cohort study of all patients diagnosed with CRC at an academic center between 1/1/2012 and 1/31/2021. Tumor testing included MMR immunohistochemistry, followed by *BRAF* V600E/*MLH1* promoter methylation testing when indicated. The intervention included a manual phase, which systematized Pathology screening and cancer genetics (CG) referral mechanisms, and an automated phase utilizing computer programming.

**Results:** A total of 249/1,541 CRC (17.38%) had MMR loss of expression and 129 (8.37%) qualified for CG evaluation. Referral was 27.58% in the original cohort and 92.1% in the intervention (p<0.001). Patients seen by CG among referred were 27.58% in the original cohort and 74.3 % in the intervention (P two-sided <0.001). The distribution of race/ethnicity among patients qualifying and referred for CG evaluation was not significantly different across cohorts. LS diagnosis increased from 0.56% (original cohort) to 1.43% (intervention). Cost per new diagnosis of LS decreased from $173,675 to $87,960 from original cohort to intervention.

**Conclusion:** Implementation of systematic case identification and referral support mechanisms significantly increased the proportion of patients undergoing genetic testing and doubled the percentage of patients diagnosed with Lynch syndrome with no referral differences across racial/ethnic groups.

## Introduction

Lynch syndrome (LS) affects 1/279 individuals (0.36% of the general population)[1] and it accounts for 3-5% of all colorectal cancers (CRC). LS is due to germline mutations in mismatch repair (MMR) genes *MLH1, MSH2, MSH6*, or *PMS2*. Surveillance and prevention measures reduce mortality[2] and benefits of diagnosing LS extend to relatives.

Over the years, criteria have been developed to identify individuals who should have genetic testing for LS. Some of these criteria select tumors to undergo pre-screening through either microsatellite instability (MSI) testing or immunohistochemistry (IHC) to evaluate MMR protein expression. Patients with MMR-deficient tumors would undergo germline testing.

About 10-15% of sporadic CRC exhibit MSI and loss of MLH1/PMS2 protein expression. These cases are secondary to hypermethylation of the *MLH1* promoter region, which distinguishes them from tumors associated with LS. Thus, individuals with *MLH1-*methylated tumors do not require genetic testing. Alternatively, CRC with loss of *MLH1* expression can be tested for the *BRAF* V600E variant. This variant is associated with *MLH1* promoter hypermethylation and thus sporadic tumors. Testing for *BRAF* V600E is not as sensitive though as only 68% of CRC with *MLH1* methylation contain *BRAF* V600E variants[3], but if present, it does rule out LS. The addition of either *MLH1* methylation or *BRAF* V600E analysis has been suggested to further restrict candidate individuals for germline testing.

While this stepwise process is effective it fails to identify about 28% of LS cases, mostly due to overly restrictive selection criteria[4]. This fact prompted the now widely adopted recommendation of testing all CRC for LS. Despite these recommendations, implementation still falls below 50% of CRC in North America[5-7]. Even when implemented, a study from four academic institutions across the US reported that only 29.5% of individuals with MMR-deficient tumors underwent genetic testing[8].

This suboptimal use of tumor testing, along with overall poor familiarity with characteristics suggestive of LS, have resulted in a significant underdiagnosis[9]. This is in spite of LS having been considered priority genomics objective of the Healthy People 2020 initiative of the US Department of Health and Human Services[10].

While racial disparities have been widely documented in cancer care, the extent of disparities in LS diagnosis is particularly alarming. In fact, African American (AA) and Hispanic (H) patients undergo genetic testing for LS at less than half the rates of Non-Hispanic Whites (NHW)[8]. Even in programs that have implemented universal tumor testing, racial/ethnic minority patients with MMR-deficient tumors have been consistently under-referred for genetic testing[8, 11].

To improve LS testing, particularly among minorities, we developed the CLEAR LS (Closed Loop Enhanced Assessment and Referral for Lynch Syndrome) intervention, a mechanism to systematically identify patients with MMR-deficient tumors and enhance referral for genetic testing. This system was eventually fully automated, dramatically decreasing manpower required to sustain it. The CLEAR LS intervention significantly increased the proportion of eligible patients undergoing genetic testing and doubled the percentage of patients diagnosed with LS with a meaningful impact on patients belonging to racial/ethnic minority groups. The costs per new diagnosis of LS were halved through this intervention.

## Methods

We conducted a pre-/post-intervention cohort study to evaluate whether implementation of CLEAR LS increased the proportion of patients appropriately evaluated, referred, and tested for LS. The study includes all patients diagnosed with CRC at Yale New Haven Hospital between 1/1/2012, and 1/31/2021. Patients diagnosed with CRC between 1/1/2012, when implementation of universal CRC testing began, and 5/31/2015, represent the original cohort. The intervention cohort includes patients diagnosed thereafter and is divided into two phases: manual 6/1/2015, through 7/31/2018, and automated from 8/1/2018, through 1/31/2021. Sex, race/ethnicity were self-reported. The study was exempt from IRB review.

### Pathology testing: Immunohistochemistry and Molecular screening

All new diagnoses of CRC underwent MMR IHC analysis using antibodies against *MLH1, PMS2, MSH2* and *MSH6* using manufacturer’s recommendations. Stained slides were interpreted for nuclear expression of each MMR protein in tumor cells. When absent MLH1/PMS2 expression was observed, the surgical Pathologist ordered subsequent *BRAF* V600E mutation analysis. The later was substituted by *MLH1* promoter methylation in 6/2018 (Figure 1). DNA was extracted from unstained, formalin fixed, paraffin-embedded tissue sections, which were micro-dissected if necessary to enrich for malignant cells (minimum malignant cell content 20%) based on corresponding H&E slide review. *BRAF* V600E was assessed using a PCR-single strand confirmation polymorphism in-house developed assay. *MLH1* methylation was performed using methylation-specific multiplex PCR after treatment of the DNA with sodium bisulfite according to manufacturer’s recommendation (Qiagen, Inc. Germantown, Maryland). All tests were interpreted by expert GI faculty in the CLIA certified pathology/molecular diagnostics laboratories.

**Figure 1.**
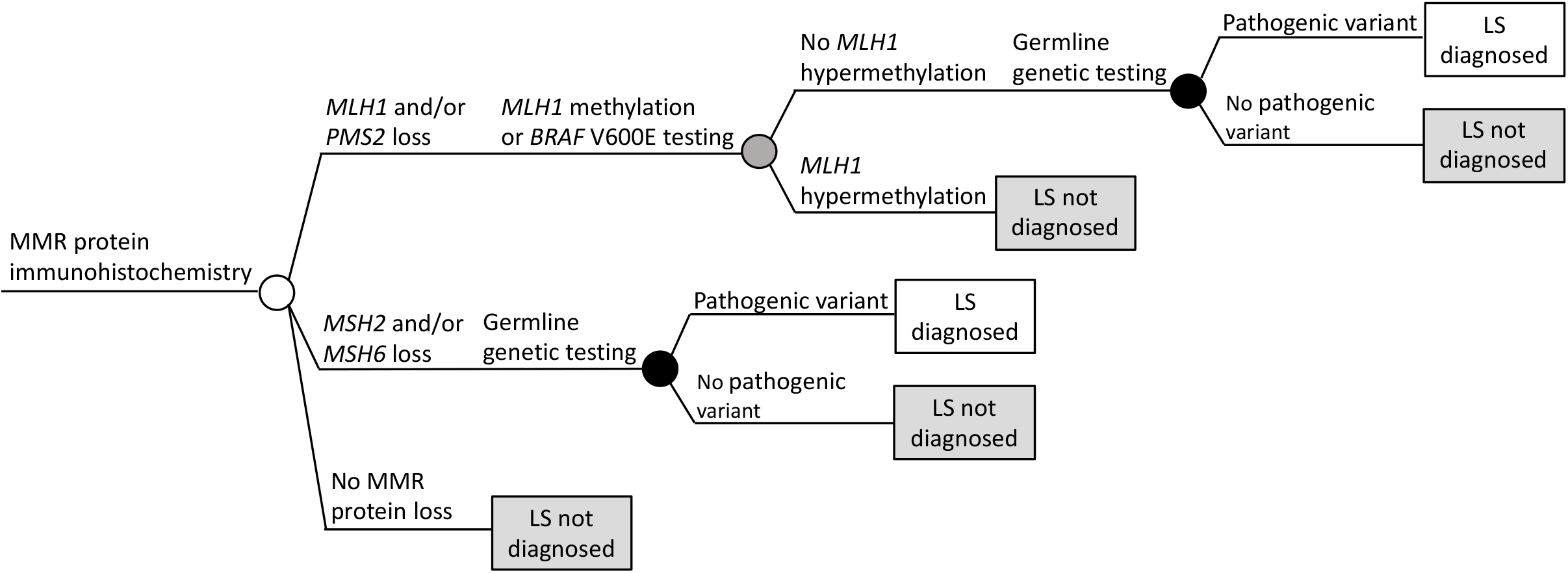
Tumor screening algorithm

### Manual Intervention Strategy

Since the implementation of universal IHC testing, pathology reports have included a paragraph recommending cancer genetics (CG) evaluation for all cases with loss of MMR expression and wild-type *BRAF* V600E or absence of *MLH1* methylation. Under this intervention, a Pathologist manually matched MMR expression with *BRAF* results and generated a weekly list of eligible cases. This list was sent through the electronic health record (EHR) to a genetic counselor at the CGPP. Two weeks after the data was shared with the CGPP, if no referral had been placed, a message was sent to the patient’s provider explaining the need for evaluation. A second and a final third message by the CGPP Director were sent 2 weeks apart when needed (Figure 2).

**Figure 2.**
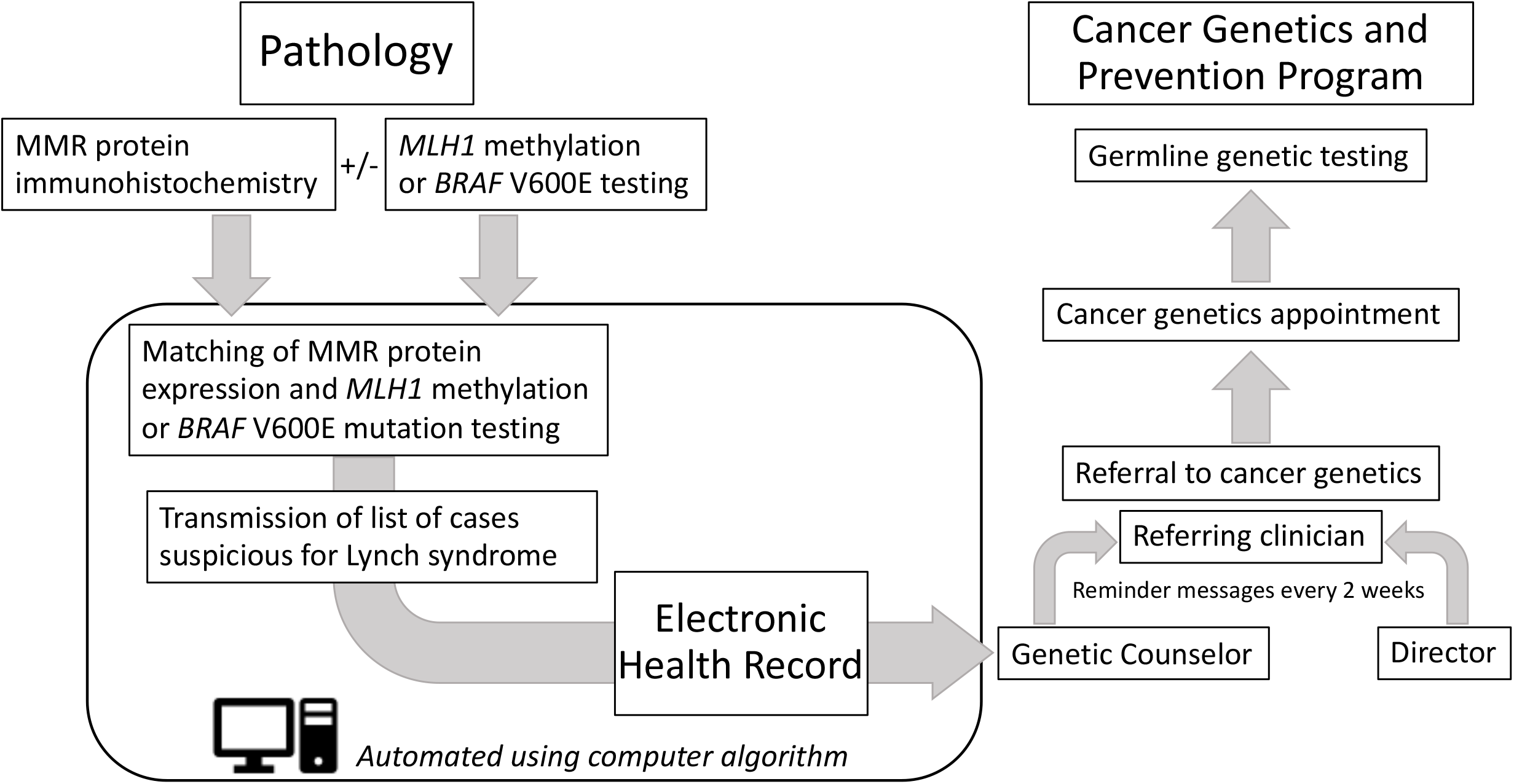
Closed Loop Enhanced Assessment and Referral for Lynch Syndrome (CLEAR LS) Intervention

### Automated Intervention Strategy

During the second part of the intervention, the entire pathology and cancer genetics communication process was automated using a Java-based algorithm and the report was automatically sent weekly to the CGPP. The web-based standalone monitoring engine called Repetitive Tasks Scheduling Engine (RTSE) was used to automate identification of patients eligible for genetic testing. Briefly, molecular tests results are reviewed and entered by Pathologists into a laboratory information system called CoPathPlus (Cerner DHT, Boston, MA). Data is stored in the underlying relational database-Adaptive Server Enterprise (SAP, Walldorf, Germany), which is accessible to RTSE via a Java Database Connectivity API. A patient report in Pathology is a complex document that includes multiple fields. These fields are stored in the database along with the formatting tags encoded using the Rich Text Format specification which makes it searchable. The RTSE runs every five minutes, collecting data from all cases signed out during that period and converting documents into plain text suitable for the indexing and search. A separate job runs weekly to retrieve cases that match criteria: lack of MMR expression and no *BRAF* V600E mutation or *MLH1* methylation. Synoptic reports are selected as a basis for filtering relevant cases. These reports are created by Pathologists for tumor resections. This job, ultimately, tabulates results and emails the report to the CGPP.

### Genetic counseling and testing

After reports are generated, a genetic counselor reviews patient lists provided by Pathology, contacts the patients’ providers, and tracks compliance with consults and testing. Genetic testing is offered through a CLIA certified laboratory.

### Cost

Institutional charges associated with billing codes for each test including: IHC, microdissection, *MLH1* methylation, *BRAF* V600E mutation, and genetic testing, were tabulated. Charges were: IHC $1,000; microdissection $765; *MLH1* methylation $200; *BRAF* V600E $430; genetic testing $3,970. For molecular testing, microdissection was required for 77% of cases.

### Statistical analysis

Continuous variables are presented as median and standard deviation, and categorical variables are presented as proportions. Patient characteristics were compared between cohorts, using two-sided tests. For continuous variables we used a non-parametric test (Kruskal–Wallis test) and for categorical variables a Fisher’s exact test. The proportion of patients that underwent each test in the algorithm was calculated and the sensitivity of the overall diagnostic system for detecting LS was assessed for the original cohort and the intervention period (combining manual and automated strategies). Statistical analyses were performed using Stata/MP 16.1.

To control for false discovery rate, reported P-values are adjusted P-values corrected using the Benjamini & Hochberg procedure in R. Adjusted P-values <0.05 were considered statistically significant.

## Results

A total of 1,541 CRC patients were diagnosed. IHC was performed on 92.34% (1,433/1,541) samples. The median age (range) was 66 (17 – 97) years (Table 1). Men were younger than women: 64 (23 – 96) years, vs. 67 (17 – 97) years, two sided P<0.001. Hispanic (57 (17 – 91) years), Asian (61 (35 – 88) years), and African American (63 (23 – 89) years) were younger than Non-Hispanic White patients (67 (13 – 97) years), two sided P<0.001.

**Table 1:** Demographic characteristics and tumor testing for identification of patients for Lynch Syndrome genetic testing. IHC: Immunohistochemistry; MMR: mismatch repair; CG: Cancer Genetics. *Four had simultaneous *BRAF* mutation).

|  |  | Total (%) | Original Cohort (%) | Intervention Cohort |  | Adjusted P-value |
| --- | --- | --- | --- | --- | --- | --- |
|  |  |  |  | Manual (%) | Automated (%) |  |
| Number of patients |  | 1541 | 354 | 783 | 404 |  |
| IHC Tested |  | 1433 (92.3) | 284 (80.23) | 747 (95.40) | 402 (99.50) | <0.001 |
| Gender |  |  |  |  |  |  |
|  | *Female* | 668/1433 (46.6) | 133/284 (46.8) | 341/747 (45.6) | 194/402 (48.26) | 0.855 |
|  | *Male* | 765/1433 (53.4) | 151/284 (53.2) | 406/747 (54.3) | 208/402 (51.74) |  |
| Median Age (SD) |  | 66 (14.13) | 64 (14.89) | 66 (13.69) | 66 (14.34) | 0.063 |
| MMR Protein Expression loss |  | 249/1433 (17.4) | 50/284 (17.61) | 130/747 (17.40) | 69/402 (17.16) | 0.986 |
|  | *PMS2/MLH1* | 180/1433(12.6) | 38/284 (13.38) | 93/747 (12.45) | 49/402 (12.19) | 0.986 |
|  | *PMS2* | 19/1433 (1.3) | 5/284 (1.76) | 9/747 (1.20) | 5/402 (1.24) |  |
|  | *MSH2/MSH6* | 34/1433 (2.4) | 4/284 (1.41) | 19/747 (2.54) | 11/402 (2.74) |  |
|  | *MSH2* | 2/1433 (0.1) | 0/284 (0.00) | 2/747 (0.27) | 0/402 (0.00) |  |
|  | *MSH6* | 14/1433 (1) | 3/284 (1.06) | 7/747 (0.94) | 4/402 (1.00) |  |
|  | *No loss of expression* | 1184/1433(82.6) | 234/284 (82.39) | 617/747 (82.60) | 333/402 (82.84) |  |
| *BRAF* V600E Mutation/*MLH1* Methylation analysis performed among the ones with loss of *MLH1/PMS2* |  | 177/199 (88.9) | 31/43 (72.09) | 94/102 (92.16) | 52/54 (96.30) | 0.004 |
| Positive *BRAF* V600E Mutation/*MLH1* Methylation among the ones with loss of *MLH1/PMS2* |  | 120/199 (60) | 21/43 (48.84) | 59/102 (57.84) | 40/54 (74) | 0.063 |
|  | *BRAF* mutation Positive only | 93/150 (62) | 21/31(67.7) | 57/ 92(62) | 19/27 (70.4) |  |
|  | *MLH1* Hypermethylation | 27/31 (87.1) | 0/0 | 2/2 (100) | 25/29 (86.2)* |  |
| CG visit indicated according to path strategy | *PMS2* or *MLH1/PMS2* def. | 79/199 (39.70) | 22/43 (51.16) | 43/102(42.16) | 14/54 (25.93) | 0.063 |
|  | *MSH2, MSH6* or *MSH2/MSH6* def. | 50/50 (100) | 7/7 (100) | 28/28 (100) | 15/15 (100) | n/a |
| Patients qualifying for CG among all CRC patients |  | 129/1541 (8.37) | 29/354 (8.19) | 71/783 (9.07) | 29/404 (7.17) | 0.756 |
| Patients qualifying for CG among all IHC tested |  | 129/1433 (9.0) | 29/284 (10.21) | 71/747 (9.50) | 29/402 (7.21) | 0.480 |
| Patients Identified for CG through the process |  | 88/129 (68.22) | 8/29 (27.59) | 56/71 (78.87) | 24/29 (82.76) | <0.001 |

### Tumor testing

IHC was performed on 92.3% (1,433/1,541) samples. Testing completion increased significantly from 80.2% (original cohort) to 95.4% (manual phase) and 99.5% (automated phase) during the intervention (P < 0.001). Among all tested, 249 (17.4%) had loss of expression of at least one MMR protein with no differences across study periods. Of the 199 with loss of *MLH1*/*PMS2* expression, 177 underwent further analysis with *MLH1* promoter methylation (n=31; 15.58%), *BRAF* mutation (n=150; 75.38%), or both (n=4; 2.26%) (Table 1). Finally, 79/199 patients qualified for CG evaluation: 51.16% (original cohort); 42.16% (manual), and 25.93% (automated) intervention cohorts (P=0.063). Fifty patients qualified because of loss of *MSH2*/*MSH6* expression (Table 1).

### Systematic intervention and Cancer Genetics referral

A total of 129/1,541 patients (8.4%) qualified for CG evaluation (9% when only considering individuals with tumor testing). The higher specificity of *MLH1* methylation vs. *BRAF* V600E mutation analysis was reflected in a decreasing proportion of individuals qualifying for genetic testing over time (Table 1). Among qualifying individuals, only 27.6% in the original cohort were identified, while 78.9% (manual) and 82.8% (automated) of the intervention cohorts were identified (Table 1). Patients not identified during the intervention included cases with pathology/molecular testing performed too late to be captured by the system.

### Cancer Genetics evaluation and Lynch Syndrome Diagnosis

Referral among patients requiring evaluation was significantly higher for the intervention cohort: 92.1% *vs*. 27.6% (P<0.0001) (Table 2). Completed CG evaluation and testing was also significantly higher for the intervention cohort: 74.3% *vs*. 27.6% (P<0.0001). Two original cohort and 13 intervention cohort patients were diagnosed with LS. An additional 4 intervention patients had previously been diagnosed with LS. The percentage of LS diagnosis among the pathology evaluated patients almost doubled from 0.8% in the original cohort to 1.54%, and more than doubled when considering all patients (0.56% *vs*. 1.4%) (Table 3). To further evaluate the impact of our intervention, we determined the difference in LS diagnosis before and after intervention. We used the LS incidence from the original cohort (0.6%) to calculate the expected LS cases to be identified from 1,187 total CRC cases. The expected number was 7, but during our intervention we diagnosed 17 patients, a 143% increase (Fisher-exact test expected vs observed P= 0.06).

**Table 2:** Identification of Lynch Syndrome patients. ^§^Four patients were already diagnosed with Lynch Syndrome and did not require further evaluation. ^¥^For the original cohort, patients seen were the same as the ones appropriately referred as there was no intervention in place to request for referrals. CG: Cancer Genetics.

|  | Original cohort (%) | Intervention cohort (%) |  |  | Adjusted P-value |
| --- | --- | --- | --- | --- | --- |
|  | Total Referred | Referral prior to Intervention | Referral after Intervention | Total referred |  |
| CG referral among requiring evaluation<sup>§</sup> | 8/29 (27.59) | 38/76 (50.00) | 32/76 (42.10) | 70/76 (92.11) | <0.0001 |
| Seen by CG among referred | 8/29* (27.59) | 32/38 (84.2) | 20/32 (62.5) | 52/70 (74.29) | <0.0001 |
| LS diagnosed among total patients seen | 2/8 (25.0) | 9/32 (28.12) | 4/20 (20.00) | 13/52 (25.0) | 1.0000 |

**Table 3:**
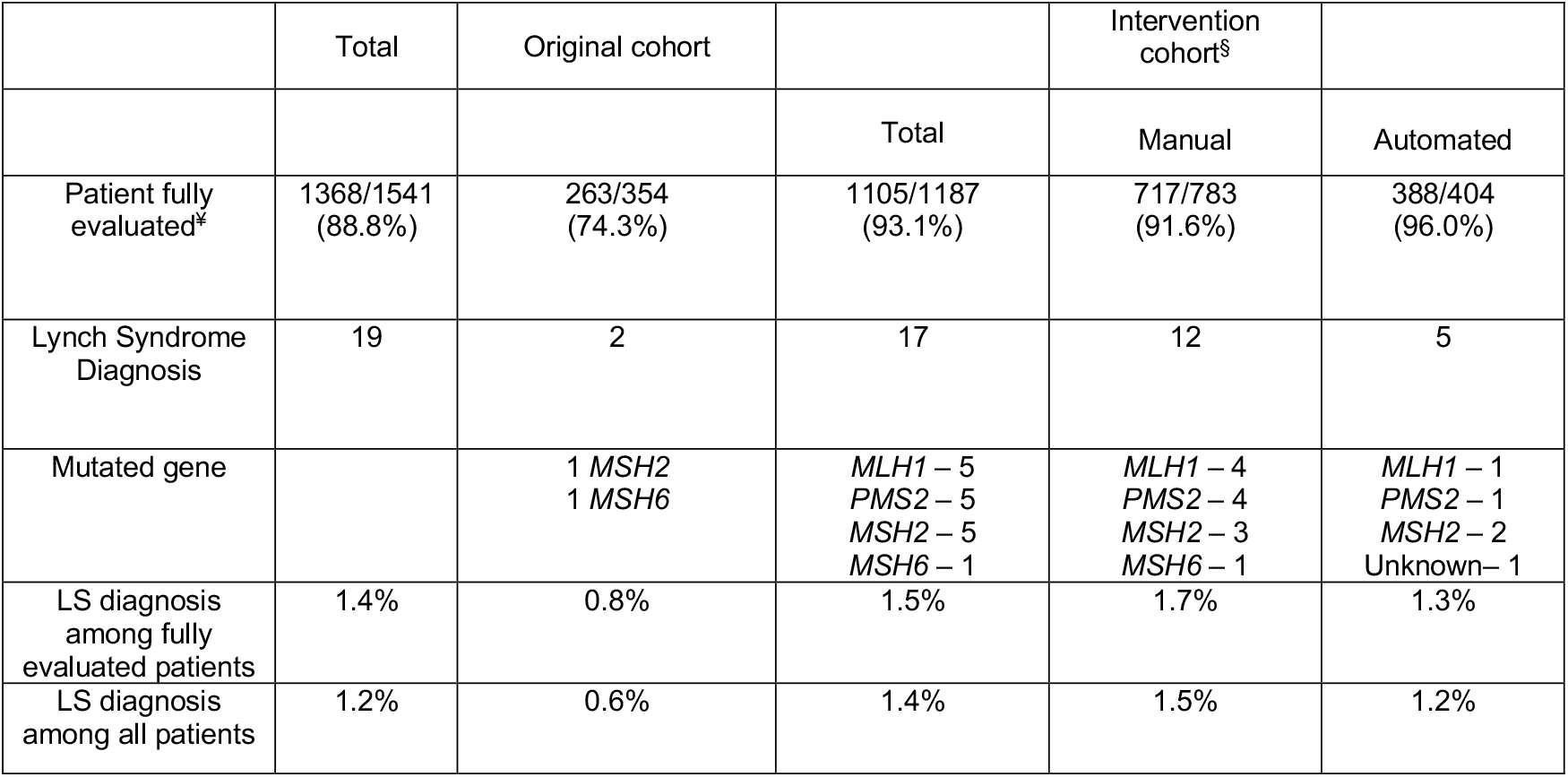
Description of patients diagnosed with Lynch Syndrome in the study period. ^§^Four patients of intervention cohort were identified to be previously diagnosed with Lynch Syndrome. ^¥^Sum of patients with confirmed normal expression of mismatch repair (MMR) protein; patients with loss of protein expression followed by *BRAF* V600E or *MLH1* promoter methylation testing if protein loss was MLH1/PMS2; and completed genetic testing in all cases when indicated. LS: Lynch syndrome

|  | Total | Original cohort |  | Intervention cohort<sup>§</sup> |  |
| --- | --- | --- | --- | --- | --- |
|  |  |  | Total | Manual | Automated |
| Patient fully evaluated<sup>‡</sup> | 1368/1541<br>(88.8%) | 263/354<br>(74.3%) | 1105/1187<br>(93.1%) | 717/783<br>(91.6%) | 388/404<br>(96.0%) |
| Lynch Syndrome Diagnosis | 19 | 2 | 17 | 12 | 5 |
| Mutated gene |  | 1 *MSH2*<br>1 *MSH6* | *MLH1* – 5<br>*PMS2* – 5<br>*MSH2* – 5<br>*MSH6* – 1 | *MLH1* – 4<br>*PMS2* – 4<br>*MSH2* – 3<br>*MSH6* – 1 | *MLH1* – 1<br>*PMS2* – 1<br>*MSH2* – 2<br>Unknown – 1 |
| LS diagnosis among fully evaluated patients | 1.4% | 0.8% | 1.5% | 1.7% | 1.3% |
| LS diagnosis among all patients | 1.2% | 0.6% | 1.4% | 1.5% | 1.2% |

Overall, sensitivity of the diagnostic system to detect LS in the original cohort was 27.6% and 51.8% during the Intervention (Figure 3). The average per-person cost of screening for LS was $981 for the original cohort and $1,260 in the intervention. However, given the increased diagnostic yield of screening with the intervention, the cost per new diagnosis of LS decreased from $173,675 in the original cohort to $87,960 in the intervention.

**Figure 3.**
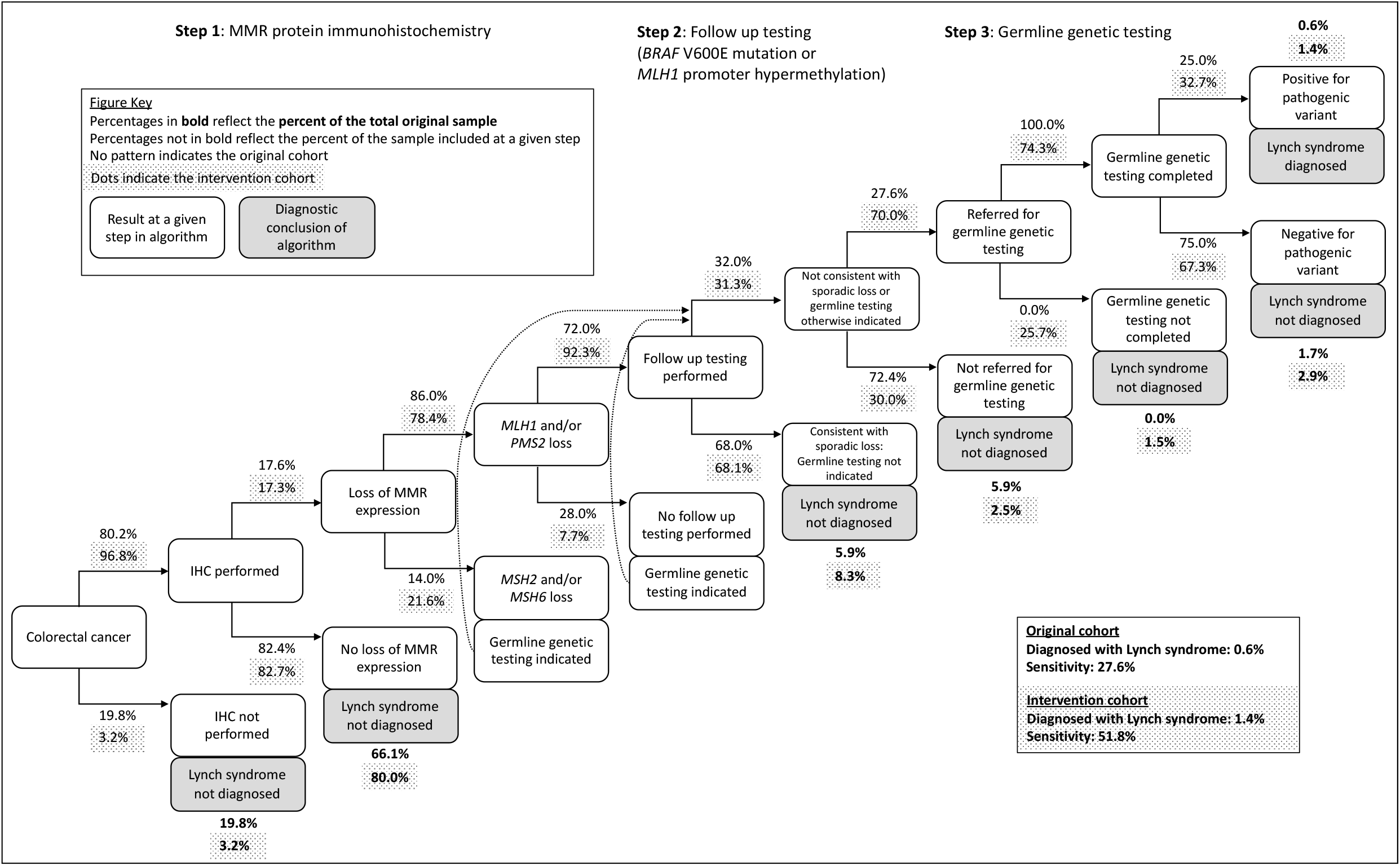
Patient screening, referral, and diagnosis in original sample and CLEAR LS intervention

### Race/Ethnicity data

Among the 1,541 patients, 79.1% were non-Hispanic Whites (NHW); 11.8%, African Americans (AA); 6.7% Hispanics (H); 2.2% Asians (A); and 0.1% were not identified (Supplementary Table). While more minority patients were appropriately referred in the intervention cohort (79.2%) than the original cohort (40%), the difference did not reach statistical significance (P=0.193) as it did for NHW patients (P<0.001) (Table 4). More minority patients were also eventually seen by CG and tested among the referred ones: 40% in the original cohort and 57.9% in the intervention (P=0.629). Significance was reached again only among NHW (P<0.001) (Table 4). There were no significant differences within the intervention cohort between NHW and the rest of the patients regarding referral (P=0.376) and eventual CG visit (P=0.193). The rest of the specific data divided by ethnic/race is summarized in the Supplementary Table.

**Table 4:**
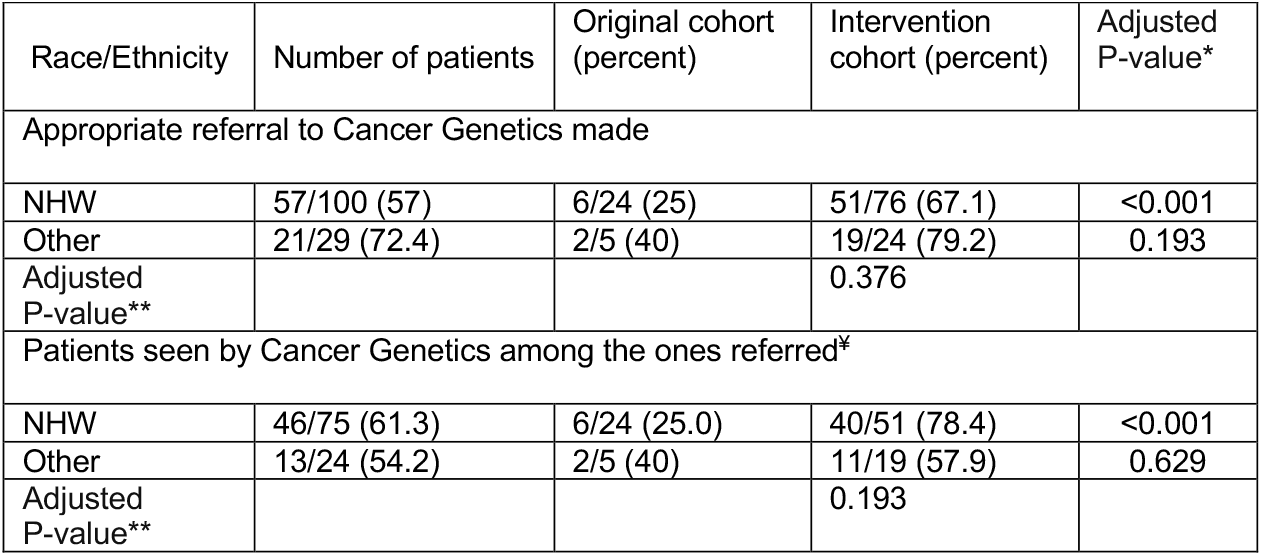
Race/ethnicity distribution of the cohorts. Non-Hispanic Whites (NHW); Other include African Americans (AA); Hispanics (H); and Asians (A). *Comparison by race/ethnic group across two cohorts. **Comparison by race/ethnic group among patients in the intervention cohort.

| Race/Ethnicity | Number of patients | Original cohort (percent) | Intervention cohort (percent) | Adjusted P-value* |
| --- | --- | --- | --- | --- |
| Appropriate referral to Cancer Genetics made |  |  |  |  |
| NHW | 57/100 (57) | 6/24 (25) | 51/76 (67.1) | <0.001 |
| Other | 21/29 (72.4) | 2/5 (40) | 19/24 (79.2) | 0.193 |
| Adjusted P-value** |  |  | 0.376 |  |
| Patients seen by Cancer Genetics among the ones referred* |  |  |  |  |
| NHW | 46/75 (61.3) | 6/24 (25.0) | 40/51 (78.4) | <0.001 |
| Other | 13/24 (54.2) | 2/5 (40) | 11/19 (57.9) | 0.629 |
| Adjusted P-value** |  |  | 0.193 |  |

## Discussion

While universal CRC tumor testing to screen for LS has been almost unanimously endorsed for years, its yield has proven modest at best[12, 13]. The reasons behind the poor performance are myriad but not surprising. At the pathology level, IHC results are usually reported later in addendum form. Then, the report must be reviewed by the clinician and if she/he decides to act (often depending on the familiarity with LS), likely it will be through a referral to CG. Then, a follow up appointment with CG must be scheduled and kept. At the visit, genetic testing is recommended, and if patients agree, they are sent to a blood draw station for sample collection or given/sent a saliva kit. The entire process involves multiple steps. Gaps in the so-called specialty referral loop are well-documented[14], and are significantly worse in minority populations[15]. The process becomes even more complex if a multi-step approach is followed using *BRAF* V600E/*MLH1* methylation analysis after identifying the absence of *MLH1*/*PMS2* expression. In this case, it is also necessary to synthesize the two tests performed at different points in time to identify patients who qualify for genetic testing. The clear advantage of this multi-step approach is that many patients can be ruled out for LS without germline testing, considerably decreasing the workload of the CG program. While feasibility of a coordinated/multidisciplinary approach in a large academic institution has already been reported[16], this requires a significant effort which can be difficult to achieve in many institutions.

The focus of the CLEAR LS was to systematize implementation and interpretation of the multi-step process while providing practical referral support mechanisms to increase genetic testing among eligible individuals. The automated active review of pathology reports, linkage to the EHR, and communication to the CG program successfully addressed the limitation posed by passive reliance on individual clinicians to return to pathology reports and appropriately interpret and act on them while also decreasing the manpower required. This approach could allow many institutions to close the referral loop and increase LS diagnosis.

Health service delivery improvements are incremental, and our intervention is no exception: while attrition still occurred at most steps in the testing and referral algorithm, we were able to significantly increase referral to CG and ultimate genetic testing and more than doubled the percentage of LS diagnosis in a universal tumor testing protocol. Furthermore, although we did not capture these additional benefits in this study, with every new LS diagnosis, other family members can be diagnosed through cascade testing, exponentially increasing the benefits of LS diagnosis.

Although we observed significant increases in completion of screening and LS diagnosis after the CLEAR LS intervention, still only about half of expected LS cases were diagnosed. It remains important to continue to evaluate the yield of universal tumor testing with respect to other approaches. Eventually, options that include more widespread germline genetic testing[17] and extending point-of-care genetic testing where cancer genetics is embedded into the oncology clinic[18] could end up identifying more individuals with LS and thus these need further consideration.

As prior studies have noted[8, 11], under-referral for CG evaluation and genetic testing disproportionally affects minorities in the US. In our series, there were no significant differences in appropriate referrals made and patients eventually seen and tested between NHW and minority populations. Thus, providers approached by the CG program responded similarly regardless of the race/ethnicity of their patients and referral bias was not perceived during the intervention. Despite this, in the intervention only 42% of AA referred completed a CG visit and were tested while all (100%) Hs and 80% of NHW who were referred completed their visits. While most patients who were not ultimately seen in CG clinic did have advanced disease, further evaluation of underlying factors impacting visit completion and targeted interventions are warranted.

While a cost-effectiveness analysis was outside the scope of this manuscript, a cost-only analysis showed that the cost per new diagnosis of LS during the intervention was about half that in the original cohort. While this cost analysis is limited, it demonstrates the impact that increasing the sensitivity and diagnostic yield of a screening system can have.

Some important limitations of this study include that this is a single center study and thus, it is unknown how the same approach would perform in other institutions. While automation of the pathology testing and CG communication results in no extra manpower effort on the Pathology side, an initial IT solution needs to be created. On the other hand, this project required some effort by the genetic counselor coordinating in-baskets to providers and tracking referrals. This effort can be offset by eliminating the need for unnecessary visits to CG by patients whose tumors were MMR-deficient due to *MLH1* methylation.

In summary, while the process of screening CRC for LS through universal tumor testing can be very useful, it is rather complex and thus, for it to be effective in the real world without challenging limited resources, a comprehensive multi-pronged approach maximizing automation turned out to be essential for its success.

## Supporting information

Patients' Race/Hispanic distribution

## Data Availability

All data produced in the present work are contained in the manuscript

## Abbreviations

AA: African Americans
CG: cancer genetics
CGPP: Cancer Genetics and Prevention Program
CLEAR LS: closed loop enhanced assessment and referral for lynch syndrome study
CRC: colorectal adenocarcinoma
HER: Electronic health record
H: Hispanics
IHC: immunohistochemistry
MSI: microsatellite instability
MMR: mismatch repair
NHW: non-Hispanic Whites
LS: Lynch syndrome
RTSE: Repetitive Tasks Scheduling Engine

