## Supplementary material for "A systems approach to enhance Lynch syndrome diagnosis through tumor testing": Patients' Race/Hispanic distribution

| Ethnicity | Number of patients | Original cohort (percent) | Intervention cohort (percent) | Comparison by ethnic group across two cohorts (p-value) |
|---|---|---|---|---|
| **Ethnic Distribution of all Patients** | | | | |
| **Total** | 1541 | 354 | 1187 | |
| **NHW** | 1219/1541 (79.1) | 287/354 (81.1) | 932/1187 (78.5) | |
| **AA** | 182/1541 (11.8) | 37/354 (10.4) | 145/1187 (12.2) | |
| **H** | 104/1541 (6.7) | 23/354 (6.5) | 81/1187 (6.8) | |
| **A** | 34/1541 (2.2) | 7/354 (2) | 27/1187 (2.3) | |
| **Other** | 2/1541 (0.1) | 0/354 (0.0) | 2/1187 (0.2) | |
| **Patients with IHC testing done** | | | | |
| **Total** | 1433/1541 (93) | 284/354 (80.2) | 1149/1187(96.8) | |
| **NHW** | 1125/1219 (92.3) | 226/287 (78.7) | 899/932 (96.6) | |
| **AA** | 175/182 (96.1) | 33/37 (89.2) | 142/145 (97.9) | |
| **H** | 97/104 (93.3) | 18/23 (78.3) | 79/81 (97.5) | |
| **A** | 34/34 (100) | 7/7 (100) | 27/27 (100) | |
| **Other** | 2/2 (100) | 0/0 (0.0) | 2/2 (100) | |
| **Patients with MMR protein loss among total tested** | | | | |
| **Total** | 249/1433 (17.4) | 50/284 (17.6) | 199/1149 (17.3) | |
| **NHW** | 210/1125 (18.7) | 44/226 (19.5) | 166/899 (18.5) | |
| **AA** | 24/175 (13.7) | 2/33 (6.1) | 22/142 (15.5) | |
| **H** | 10/97 (10.3) | 3/18 (16.7) | 7/79 (8.9) | |
| **A** | 5/34 (14.7) | 1/7 (14.3) | 4/27 (14.9) | |
| **Other** | 0/2 | 0/0 | 0/2 | |
| **Cancer genetics visit indicated among IHC tested** | | | | |
| **Total** | 129/1433 (9.0) | 29/284 (10.2) | 100/1149 (8.7) | |
| **NHW** | 100/1125 (8.9) | 24/226 (10.6) | 76/899 (8.4) | |
| **AA** | 16/175 (9.1) | 2/33 (6.1) | 14/142 (9.9) | |
| **H** | 9/97 (9.3) | 2/18 (11.1) | 7/79 (8.9) | |
| **A** | 4/34 (11.8) | 1/7 (14.3) | 3/27 (11.1) | |
| **Other** | 0/2 (0) | 0 | 0/2 (0) | |
| **Appropriate referral to Cancer Genetics made** | | | | |
| **Total** | 78/129 (60.5) | 8/29 (27.6) | 70/100 (70) | |
| **NHW** | 57/100 (57) | 6/24 (25) | 51/76 (67.1) | |
| **AA** | 12/16 (75) | 0/2 (0.0) | 12/14 (85.7) | |
| **H** | 6/9 (66.7) | 1/2 (50) | 5/7 (71.4) | |
| **A** | 3/4 (75) | 1/1 (100) | 2/3 (66.7) | |
| **Other** | 0/0 (0) | 0/0 (0) | 0/0 (0) | |
| **Patients seen by Cancer Genetics among the ones referred[¥]** | | | | |
| **Total** | 59/99 (59.6) | 8/29 (27.6) | 51/70 (72.9) | |
| **NHW** | 46/75 (61.3) | 6/24 (25.0) | 40/51 (78.4) | |
| **AA** | 5/14 (35.7) | 0/2 (0) | 5/12 (41.7) | |
| **H** | 6/7 (85.7) | 1/2 (50) | 5/5 (100) | |
| **A** | 2/3 (66.7) | 1/1 (100) | 1/2 (50) | |
| **Other** | 0/0 (0) | 0/0 (0) | 0/0 (0) | |
| **Patients diagnosed with Lynch syndrome among entire cohort** | | | | |
| **Total** | 19/1541 (1.2) | 2/354 (0.56) | 17/1187 (1.4) | |
| **NHW** | 12/1219/ (1) | 1/287 (0.3) | 11/932 (1.2) | |
| **AA** | 3/182 (1.6) | 0/37 (0) | 3/145 (2.1) | |
| **H** | 3/104 (2.9) | 1/23 (4.3) | 2/81 (2.5) | |
| **A** | 1/34 (2.9) | 0/7 (0) | 1/27 (5.9) | |
| **Other** | 0/2 (0) | 0/0 (0) | 0/2 (0) | |

**Supplementary table.** Non-Hispanic Whites (NHW); African Americans (AA); Hispanics (H); Asians (A)
